# Shifting research priorities in maternal and child health in the COVID-19 pandemic era in India: a renewed focus on systems strengthening

**DOI:** 10.1101/2021.02.28.21252648

**Authors:** Kayur Mehta, Sanjay Zodpey, Preetika Banerjee, Stephanie L. Pocius, Baldeep K. Dhaliwal, Andrea DeLuca, Sangeeta Das Bhattacharya, Shailendra Hedge, Paramita Sengupta, Madhu Gupta, Anita Shet

**Author notes:** Address correspondence to: Dr. Anita Shet, MD, PhD., Director, Maternal and Child Health Center India, International Vaccine Access Center, 415 N Washington Street 5th Floor, Baltimore, MD 21231.

## Abstract

**Background:** The remarkable progress seen in maternal and child health (MCH) in India over the past two decades has been impacted by setbacks from the COVID-19 pandemic. We aimed to undertake a rapid assessment to identify key priorities for public health research in MCH in India within the context and aftermath of the COVID-19 pandemic.

**Methods:** A web-based survey was developed to identify top research priorities in MCH. It consisted of 26 questions on six broad domains: vaccine preventable diseases, outbreak preparedness, primary healthcare integration, maternal health, neonatal health, and infectious diseases. Key stakeholders were invited to participate between September and November 2020. Participants assigned importance on a 5-point Likert scale, and assigned overall ranks to each sub-domain research priority. Descriptive statistics were used to examine Likert scale responses, and a ranking analysis was done to obtain an “average ranking score” and identify the top research priority under each domain.

**Results:** Amongst the 84 respondents, 37% were public-health researchers, 25% healthcare providers, 20% academic faculty and 13% were policy makers. Across the six domains, most respondents considered conducting research on systems strengthening as extremely important. The highest ranked research priorities were strengthening the public sector workforce (vaccine preventable diseases), enhancing public-health surveillance networks (outbreak preparedness), nutrition support through community workers (primary care integration), encouraging at least 4-8 antenatal visits (maternal health), neonatal resuscitation to reduce birth asphyxia (neonatal health) and pediatric and maternal screening and treatment of tuberculosis (infectious diseases). Common themes identified through open-ended questions were also systems strengthening priorities across domains.

**Conclusions:** The overall focus for research priorities in MCH in India during the COVID-19 pandemic is on strengthening existing services and service delivery, rather than novel research. Our results highlight pivotal steps within the roadmap for advancing and sustaining maternal and child health gains during the ongoing COVID-19 pandemic and beyond.

## Background

Maternal and child health (MCH) outcomes have been key public health issues in developing countries including India. While the past 25 years have witnessed a concerted global effort to improve MCH in low-and-middle-income countries (LMICs), more progress is needed to meet the targets laid out in the Sustainable Development Goals (SDGs). In India, key MCH indicators remain concerning, as evidenced by partially released results from the National Family Health Survey-5 (NFHS-5), which indicate worsening of maternal and child nutrition indicators in several states (1). There is an urgent need to identify targeted approaches to improve the state of MCH in India.

While MCH programs in India are weighed down by wide inequities (1, 2), these challenges have been amplified by the ongoing COVID-19 pandemic. In addition to the direct effects of the disease, the indirect effects of the pandemic have been devastating. Limitations in the availability of skilled health workers and health system access barriers have led to lower coverage of antenatal and postnatal care services, and routine childhood immunization services, especially in rural India (3, 4). A recent modelling paper estimated that the disruption and decreased utilization of routine health services and reduced access to food during the pandemic could lead to an estimated 9.8–44.7% increase in under-5 child deaths per month, and an 8.3–38.6% increase in maternal deaths per month in LMIC settings (5).

Health research has been noted to be an essential tool for improving health outcomes in LMICs (6). Research prioritization during the COVID-19 pandemic using a methodical and inclusive process could guide researchers, policy makers and funding agencies in making informed decisions and improving outcomes. We aimed to undertake a rapid assessment to identify key priorities for public health research in MCH in India within the context of the COVID-19 pandemic, by surveying key stakeholders and experts in India.

## Methods

### Survey questionnaire and design

A survey to assess research needs in MCH was developed by researchers at the Johns Hopkins Maternal and Child Health Center in India and key Indian institutions. Specific priority research domains were identified through a review of previous survey results (2), and literature on global priorities in maternal newborn and child health. The survey consisted of 26 questions which focused on the following six broad domains: vaccine preventable diseases, outbreak preparedness, primary healthcare integration, maternal health, neonatal health, and infectious diseases. Under each domain, participants were requested to assign importance to each sub-domain research priority on a 5-point Likert scale, and assign overall ranks to each sub-domain research priority. Open-ended questions were also included to capture other aspects of research priorities. In each domain, participants had the option to nominate a different research topic not included in the prepared list; these could be ranked by importance in the same manner. The study was reviewed by the JHSPH Institutional Review Board (IRB) and determined to be exempt from human subjects research oversight.

### Participants

Key stakeholders working in the field of MCH in India were identified through desk reviews, web-based searches, dissemination in professional networks such as the Indian Association of Preventive and Social Medicine, Indian Public Health Association, Indian Academy of Pediatrics and the Federation of Obstetric and Gynaecological Societies of India, and through collaborators.

### Dissemination of the survey

The survey was administered online using the software platform Qualtrics (Qualtrics, Provo, UT) between September and November 2020. It was disseminated via email, text messages and social media platforms such as WhatsApp. No personal data were collected, although respondents could choose to share their contact information for future surveys.

### Data analysis

Descriptive statistics, expressed as frequencies and proportions, were used to examine respondent demographics and Likert scale responses. The Likert scale for each sub-domain research priority was analyzed by tabulating responses based on importance level ascribed. A ranking analysis was conducted using weights that were inversely related to the raw rank from the survey, to obtain an “average ranking score”, which helped identify the top research priorities in each research domain (7). Responses to the open-ended items were assessed by two authors (SLP and KM). Comments were read independently, and summarized following discussion according to the principles of thematic analysis.

## Results

### Participant characteristics

The online survey was completed by 84 respondents from 15 states in India. Male respondents constituted 58%, and professional backgrounds were distributed as follows: public health researchers (37%), healthcare providers (25%), academic teaching faculty (20%) and policy makers (13%). An almost equal number of the respondents belonged to non-governmental organizations/private organizations or governmental organizations, and over 65% had work experience of over 10 years in their primary area of expertise. The most common primary areas of work included child health (20%), infectious diseases (17%), health systems and policy (14%), biostatistics and epidemiology (14%), and maternal health (8%) (Table 1).

**Table 1.** Characteristics of survey respondents

|  |  |
| --- | --- |
| **Gender** | **N=84 (n,%)** |
| Male | 49 (58) |
| Female | 35 (42) |
| **Profession** | **N = 84 (n, %)** |
| Public health researcher | 31 (36.9) |
| Healthcare worker | 21 (25.0) |
| Academia member or teacher | 17 (20.2) |
| Policymaker | 11 (13.1) |
| Other | 4 (4.8) |
| **Primary organization type** | **N = 83 (n,%)<sup>†</sup>** |
| Non-governmental organization (NGO)/<br>private organization | 31 (37.3) |
| Government / Ministry of Health | 29 (34.9) |
| University | 13 (15.6) |
| WHO/UN agencies | 6 (7.2) |
| Other | 5 (6.0) |
| **Primary area of work** | **N = 84 (n, %)** |
| Child health | 17 (20.2) |
| Infectious disease | 14 (16.6) |
| Health systems and policy | 12 (14.2) |
| Biostatistics and epidemiology | 12 (14.2) |
| Maternal health | 7 (8.3) |
| Health economics | 3 (3.5) |
| Nutrition | 3 (3.5) |
| Rural/Tribal health outreach | 2 (2.3) |
| Public health planning | 2 (2.3) |
| Other | 16 (19.0) |
| **Number of years worked in primary area** | **N = 82 (n, %)<sup>††</sup>** |
| 1-5 | 11 (13.4) |
| 5-10 | 16 (19.6) |
| 10-20 | 27 (32.9) |
| > 20 | 28 (34.1) |

<sup>†</sup>Data were missing for 1 participant.
<sup>††</sup>Data were missing for 2 participants.

### Importance of research topics on the Likert scale

Across the six domains identified in the survey, most respondents characterised conducting research on systems strengthening priorities as extremely important (Figure 1). In the vaccine preventable diseases domain, over 50% of the respondents considered enhancing vaccination rates among the urban poor, strengthening the public sector workforce and improving vaccination coverage as extremely important. In the outbreak preparedness domain, the proportion who considered the sub-domain research priorities as extremely important ranged from 48% (conducting COVID-19 research for MCH sub-populations) to 76% (enhancing public health surveillance networks). In the primary care integration domain, over 45% of the respondents considered all sub-domain research priorities as extremely important, and 63% felt research on nutrition education support through community health workers was extremely important. In the maternal health domain, systems strengthening was considered extremely important, with 64% considering research on improving facility-based safe delivery outcomes as extremely important. In the neonatal health domain, system strengthening priorities were similarly deemed extremely important, with over 75% considering improving the quality of care during labor and birth, and 67% considering measures to reduce birth asphyxia and improve the initiation of breastfeeding in the hospital as extremely important. In the infectious diseases domain, 63% and 45% of the respondents considered conducting research on diarrhoeal diseases and COVID-19 respectively, extremely important.

**Figure 1:**
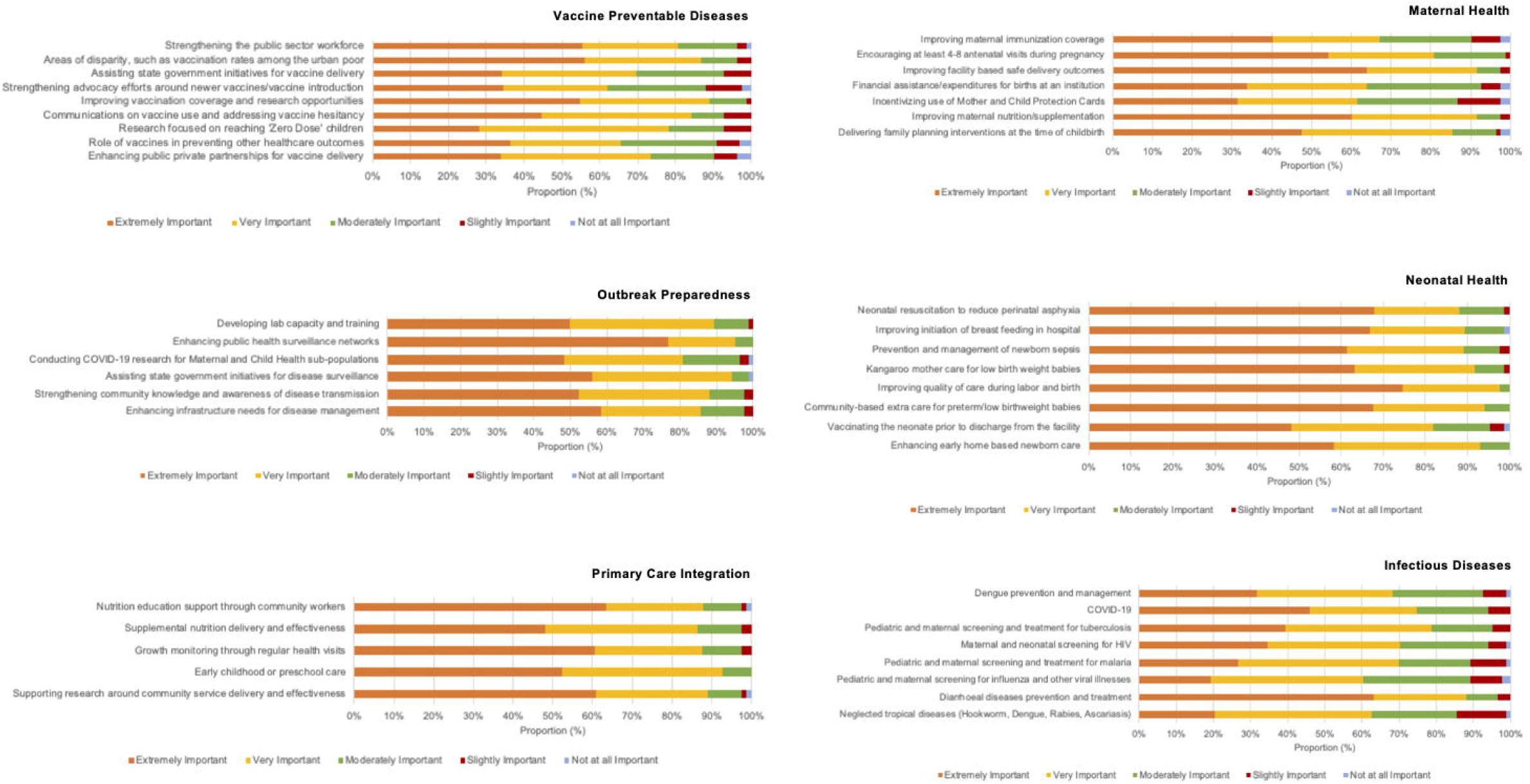
Level of importance assigned to individual sub-domain research priorities by respondents across six domains of maternal and child health (n=84).

### Overall ranking of research priorities

The highest ranked research priorities identified across domains were strengthening the public sector workforce (vaccine preventable diseases), enhancing public health surveillance networks (outbreak preparedness), nutrition support through community workers (primary care integration), encouraging at least 4-8 antenatal visits during pregnancy (maternal health), neonatal resuscitation to reduce birth asphyxia (neonatal health), and pediatric and maternal screening and treatment of tuberculosis (infectious diseases) (Table 2). While the Likert scale importance assignment and overall ranking exercises were concurrent across most domains, in the infectious diseases domain, a large proportion of respondents felt research on diarrhoeal disease prevention and treatment was extremely important; the ranking exercise, however, yielded pediatric and maternal screening and treatment for tuberculosis as the top priority.

**Table 2:**
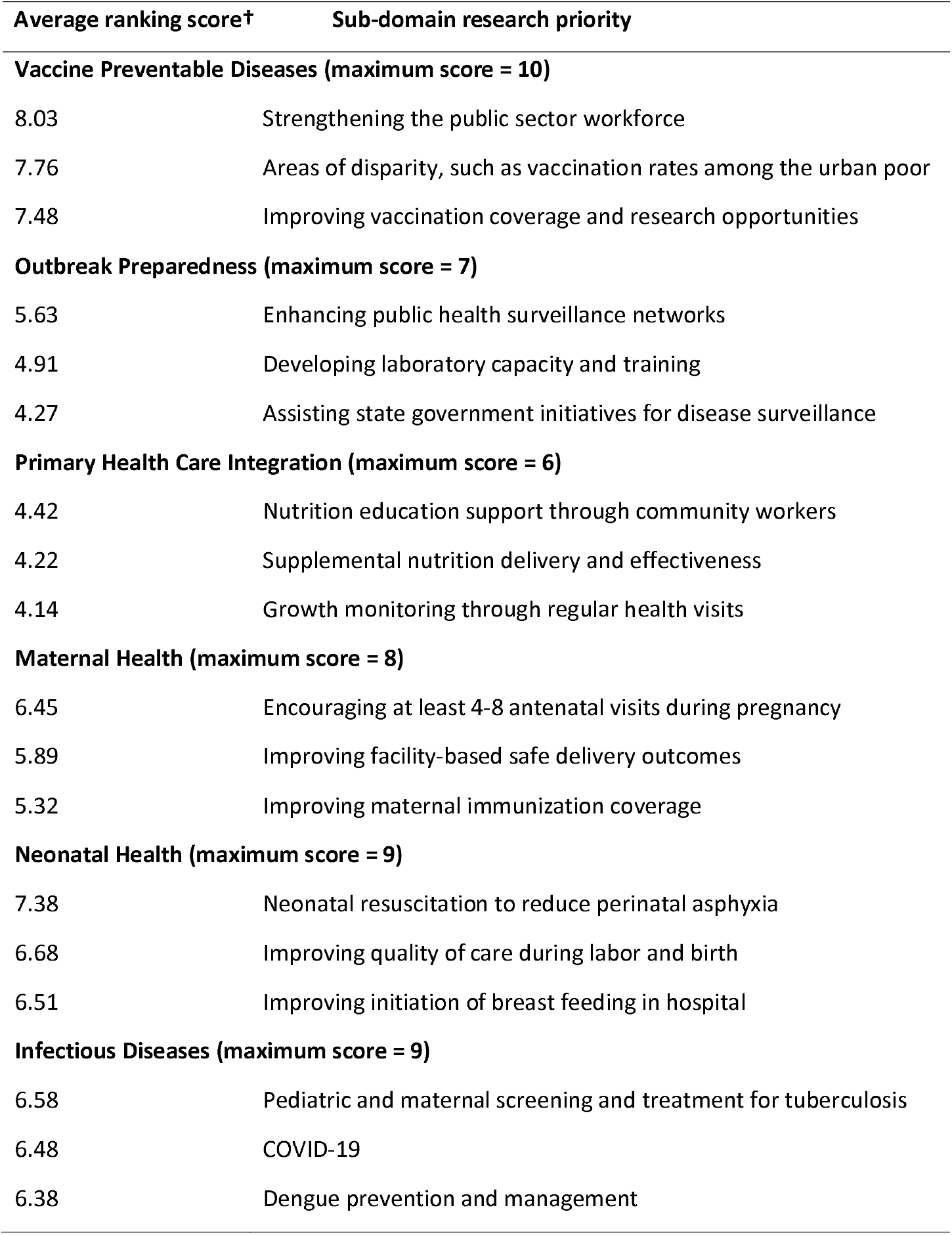
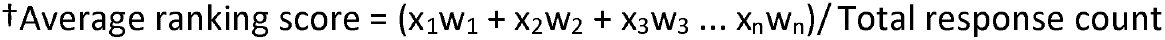
Top 3 ranked research priorities across each domain, by average ranking score

| Average ranking score† | Sub-domain research priority |
| --- | --- |
| **Vaccine Preventable Diseases (maximum score = 10)** |  |
| 8.03 | Strengthening the public sector workforce |
| 7.76 | Areas of disparity, such as vaccination rates among the urban poor |
| 7.48 | Improving vaccination coverage and research opportunities |
| **Outbreak Preparedness (maximum score = 7)** |  |
| 5.63 | Enhancing public health surveillance networks |
| 4.91 | Developing laboratory capacity and training |
| 4.27 | Assisting state government initiatives for disease surveillance |
| **Primary Health Care Integration (maximum score = 6)** |  |
| 4.42 | Nutrition education support through community workers |
| 4.22 | Supplemental nutrition delivery and effectiveness |
| 4.14 | Growth monitoring through regular health visits |
| **Maternal Health (maximum score = 8)** |  |
| 6.45 | Encouraging at least 4-8 antenatal visits during pregnancy |
| 5.89 | Improving facility-based safe delivery outcomes |
| 5.32 | Improving maternal immunization coverage |
| **Neonatal Health (maximum score = 9)** |  |
| 7.38 | Neonatal resuscitation to reduce perinatal asphyxia |
| 6.68 | Improving quality of care during labor and birth |
| 6.51 | Improving initiation of breast feeding in hospital |
| **Infectious Diseases (maximum score = 9)** |  |
| 6.58 | Pediatric and maternal screening and treatment for tuberculosis |
| 6.48 | COVID-19 |
| 6.38 | Dengue prevention and management |

†Average ranking score = $(x_1w_1 + x_2w_2 + x_3w_3 \dots x_nw_n) / \text{Total response count}$

### Other research priorities identified

Several additional research topics were also suggested by the respondents across the six domains. Common themes identified were systems strengthening priorities like enhancing vaccine programme surveillance, empowering communities and community health workers, enhancing the quality of antenatal and postnatal care, and health systems strengthening for the infectious diseases domain. Novel research approaches for COVID-19 related research, and the use of technology like telemedicine, geographic information systems and artificial intelligence for the effective delivery of services across domains were also suggested, but these were not considered to be as important as system strengthening priorities during the ongoing COVID-19 pandemic.

## Discussion

The key theme that emerged from this study seeking to identify MCH public health research priorities in India within the context of an unprecedented pandemic was the importance of systems strengthening rather than undertaking novel research. The findings of this exercise appeared to signal a shift from those of recent priority setting exercises in MCH, both from India and globally, which were conducted prior to the COVID-19 pandemic. Wazny et al employed the Child Health and Nutrition Research Initiative (CHNRI) method for research priority setting for child health research in India for the period between 2016-2025, with research ideas crowd-sourced from a network of experts across India (8). The majority of the top research priorities identified were related to the development of cost-effective interventions and their implementation, impact evaluations, improving data quality and monitoring of existing programs, and improving the management of morbidities (8). An Indian Council of Medical Research study to identify top research priorities in MCH found that the delivery domain of research which included implementation research, constituted about 70% of the top ten research options (9). A global exercise to characterize maternal and perinatal health priorities beyond 2015 identified research priorities that were mostly related to implementation research and the innovative development of simplified, cost-effective adaptations of existing interventions (10). These studies also identified areas for novel or “discovery” research in MCH, such as the development of innovative point of care diagnostics and technological solutions for acute maternal, neonatal and childhood morbidities, which were ranked high in priority lists. These studies employed the CHNRI method, which offers a detailed, systematic algorithm for the identification of research priorities that pools individual scorings of research options based on five weighted criteria, leveraging the collective wisdom of various stakeholders (11). This method involves several stages, which could take months to complete. Our study, positioned to fulfill the need for a rapid assessment during the COVID-19 pandemic found that prioritizing research endeavors in MCH to focus on strengthening existing service delivery was most important, and this theme was cross-cutting across the six domains.

Epidemics in the past have demonstrated that they are highly disruptive to existing health systems. Studies conducted during the West African Ebola outbreak of 2014 have shown that the indirect effects of the pandemic on maternal health were severe, resulting in decreased utilization of antepartum, intrapartum, and postpartum care (12). Significant decreases in coverage for most vaccines were also observed (13). A prospective observational study at a tertiary hospital in India reported a reduction in institutional deliveries, increase in pregnancies with complications, and increase in intensive care unit admissions between the months of April-August 2020 (14). Only a third of women had adequate antenatal care visits during this period (14). Considering the threat posed by the pandemic on previous gains in MCH, focusing interventions and policies around the identified priority areas would be crucial to steadying the health outcomes of mothers and children.

An unintended consequence of lockdowns and movement restrictions resulting from the COVID-19 pandemic in LMICs like India is their influence on ongoing transmissions of other non-COVID related infectious diseases. India reports the highest burden of tuberculosis globally, accounting for a quarter of the 10 million global cases and 1.4 million deaths annually (15). A recent modelling analysis of the impact on COVID-19 related disruptions in tuberculosis services in India showed that even temporary disruptions could cause long term increases in tuberculosis incidence and mortality (16). In our study, under the infectious diseases domain, conducting research on tuberculosis was ranked as the top research priority, highlighting the fact that tuberculosis continues to remain highly relevant in the Indian setting, despite the emergence of COVID-19. Further, a majority of respondents also felt that conducting research on diarrhoeal diseases was extremely important, emphasizing the need to continue to focus efforts on the prevention and treatment of diarrhoeal diseases in the Indian context. The overall ranking for diarrhoeal diseases, however, occurred below that of tuberculosis, COVID-19, and dengue infections, underscoring the changing patterns of high-burden infectious diseases in India. The identification of non-COVID-19 infectious diseases as leading research priorities in the infectious diseases domain in a survey administered during the unprecedented pandemic highlights the collective opinion of the respondents on the need to sustain research focus on other non-COVID-19 infectious diseases as well.

While there are useful lessons learned from these findings, the study limitations are worth noting. The survey was distributed within a short timeline between September - November 2020, and the results are limited by the small sample size and possible selection bias. Since this was a rapid assessment, we did not use qualitative methods to capture perspectives, and priorities were identified solely based on overall ranking and Likert scales. Further, the survey did not explore priorities for chronic and non-communicable diseases which have also witnessed an escalating burden in India in recent years. Diagnosis and treatment for such diseases have likely been affected by COVID-19 as well and warrant further research. Despite these limitations, our study has been able to perceive a shift in research priorities for MCH towards systems strengthening during this unprecedented pandemic, and can serve as a valuable resource for donors, researchers and other stakeholders engaged in maternal and child health in India.

The COVID-19 pandemic and its response has significantly challenged health systems globally. It has exposed glaring loopholes in health systems, and has demonstrated that countries’ response to pandemics is ultimately dependent on the resilience of their health systems (17). Countries such as New Zealand, Sri Lanka, Taiwan and South Korea have been praised for their handling of the COVID-19 crisis; they were able to do so efficiently by investing heavily in their public health systems to improve testing, contact tracing and outreach measures early in the course of the pandemic (18). Several countries from each of the six WHO regions are demonstrating that investing in stronger health systems is crucial in responding to COVID-19, delivering the best returns on investment and making health for all a reality (19). In India, the private sector has remained a major player in healthcare delivery for several decades (20), but the ongoing pandemic has highlighted the indisputable role of the public health sector. The public health sector has not only shouldered a vast majority of preventive and outreach services, but also clinical care, with a substantial proportion of critical COVID-19 cases being treated by public health sector services (21). States with robust, well-staffed and equipped public health systems have been more successful in containing the pandemic, and as a result, public health has reached the forefront of India’s pandemic response and post-pandemic discourse. India’s recent budget has promised substantial increases in healthcare allocation and investment on public health infrastructure (22), and these are welcome steps.

In conclusion, the results of this rapid assessment suggest that in order to build a sustainable future for maternal and child health during and post COVID-19 in India, national authorities should prioritize investments and develop frameworks to strengthen existing public health systems. Governmental investments in health research should be measured against defined benchmarks (23), and central and state governments should invest in building research capacity by developing a local workforce of well-trained and motivated researchers. Stronger collaborations between researchers and policymakers will ensure that research is taken outside the academic institutions and into public health programmes. Further, national research networks could be established to coordinate research efforts by fostering collaboration and information exchange between academic institutions, governmental and non-governmental organizations. Such partnerships could help in optimally tapping the individual sectors’ strengths and contribute to the journey towards achieving the Sustainable Developmental Goals and Universal Health Coverage for all by 2030. General recommendations for establishing research priorities in maternal and child health in India are summarized in Figure 2. While our results highlight pivotal steps to the roadmap for sustaining and advancing maternal and child health during and after the COVID-19 pandemic in the Indian setting, these learnings are relevant to other emerging global economies as well.

**Figure 2:**
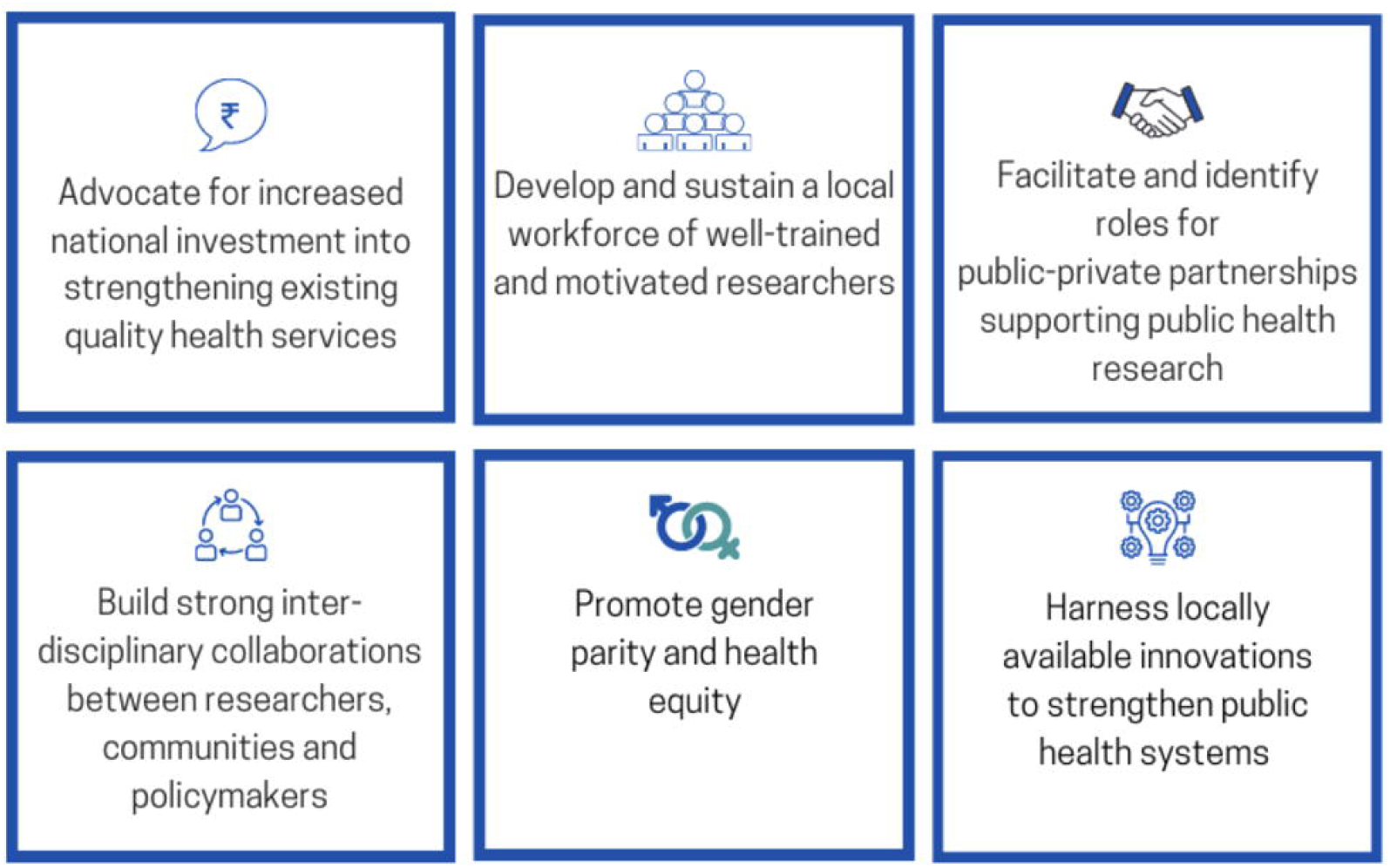
Recommendations for establishing research priorities in maternal and child health.

Where, w = weight of ranked position; x = response count for each rank. Weight of the ranked position assigned in reverse, with the respondent’s most preferred choice receiving the largest weight, which was based on the number of subdomains within each domain.

## Data Availability

All data have been referred to in the manuscript.

## Author contibutions

AS and AD conceptualized the study, and PB, BKD, SDB and KM designed the study; SZ, MG, SH, PS contributed to dissemination and data acquisition; SLP, PB and KM did the data analysis; KM wrote the initial manuscript draft, and all authors critically reviewed, provided input, and approved the final manuscript as submitted.

## Acknowledgements

The authors acknowledge funding provided by the Johns Hopkins Maternal and Child Health Center - India, and support from the Indian Association of Preventive and Social Medicine (IAPSM).

